# Assessment of *ATXN2* Repeat Expansion Length and Risk of ALS: A Meta-Analysis

**DOI:** 10.64898/2026.01.09.26343811

**Authors:** Allison A. Dilliott, Alfredo Iacoangeli, Project MinE ALS Sequencing Consortium, Ammar Al-Chalabi, Ahmad Al Khleifat, Sali M.K. Farhan

**Affiliations:** The Montreal Neurological Institute-Hospital, Montreal, QC, Canada; Department of Biostatistics and Health Informatics, King’s College London SE5 8AF, UK; Department of Basic and Clinical Neuroscience, Institute of Psychiatry, Psychology and Neuroscience, King’s College London, London SE5 8AF, UK; Perron Institute for Neurological and Translational Science, Ground RR Block QE II Medical Centre Ralph & Patricia Sarich Neuroscience Building, 8 Verdun St, Nedlands, WA 6009, Australia; NIHR Maudsley Biomedical Research Centre (BRC), South London and Maudsley NHS Foundation Trust, 16 De Crespigny Park, London SE5 8AF, UK; Department of Human Genetics, McGill University, Montreal, QC, Canada; Department of Neurology and Neurosurgery, McGill University, Montreal, QC, Canada

**Keywords:** amyotrophic lateral sclerosis (ALS), *ATXN2*, meta-analysis, repeat expansion, risk association

## Abstract

*ATXN2* expansions of ≥33 CAG-repeats are associated with spinocerebellar ataxia type 2, while “intermediate” length expansions have been associated with amyotrophic lateral sclerosis (ALS). Yet, no consensus is established regarding the lengths that define a true association with ALS risk, with recent studies debating between a lower limit of ≥29 or ≥31 repeats. Here, we assessed the risk of ALS imparted by various *ATXN2* repeat lengths to establish an accepted lower limit of repeats that impart risk of disease in the largest meta-analysis to-date.

We identified 19 studies with carrier counts of expansions ranging from 24 to ≥34 repeats in cohorts of individuals with ALS and controls that we meta-analysed with the *ATXN2* repeat lengths of the large-scale Project MinE ALS Consortium dataset (total individuals with ALS = 19202; total controls = 22177) and determined a lower limit of 30 repeats defining significant ALS risk. These findings were validated with a secondary assessment of the individuals with ALS captured within the meta-analysis using the gnomAD short tandem repeat dataset as a proxy control cohort. We also applied our defined *ATXN2* repeat risk threshold to explore relationships with ALS clinical outcomes. While we did not observe a significant relationship between *ATXN2* repeat lengths and age of ALS onset, we did identify a significant inverse correlation between *ATXN2* repeat lengths as a continuous metric and duration of disease and found that individuals with ALS carrying the risk variant allele of ≥30 repeats had significantly shorter times to diagnosis than those without the repeat expansion.

Our comprehensive analyses propose a lower-limit threshold of ≥30 *ATXN2* trinucleotide repeats in length defining true ALS risk. These findings are imperative for allowing improved accuracy in risk interpretation and guidance for patients and their families, particularly as clinical genetic testing efforts continue to expand and there becomes increased need for guiding targeted clinical trial inclusion criteria. Our results may also aid in future analyses assessing *ATXN2* pathogenic mechanisms and therapeutic strategies.

## Introduction

*ATXN2* CAG trinucleotide repeat expansions of ≥33 repeats are well-established to be causative for spinocerebellar ataxia type 2^1^; however, subsequent research also demonstrated that there is an association between “intermediate length” repeat expansions in the gene and amyotrophic lateral sclerosis (ALS)^2^. Although the biological mechanism by which the repeat expansions contribute to ALS remains largely unknown, multiple independent studies have reproduced the association. Yet the thresholds used to define an “intermediate” repeat length that is relevant to ALS largely vary across analyses, often with limited explanation as to how the limits were selected.

Large-scale efforts have been set forth to determine a more specific range of repeat expansion lengths that may confer ALS risk. Many studies have presented the lower limit of repeats defining ALS risk as *ATXN2* expansions ≥29 repeats in length^3–6^, including the previous largest meta-analysis to-date that also described that the risk of ALS increased exponentially with the length of alleles between 29 and 32 repeats^7^. While other studies have identified or deferred to a lower limit of ≥31 repeats as defining ALS risk^8–15^. These differing conclusions, along with the many other studies presenting additional potential lower limits of risk-defining repeats lengths, present challenges with investigating the functional mechanisms by which the trinucleotide repeats may impart disease risk and complicate clinical genetic testing efforts.

Here, we assessed the risk of ALS imparted by various *ATXN2* repeat lengths to establish an accepted lower limit that indeed impart risk of disease. We investigated associations with repeats of varying lengths and ALS using the large-scale Project MinE ALS Consortium dataset (6210 individuals with ALS and 2391 controls)^16^, followed by validation of our findings using meta-analyses with previous large-scale assessments of *ATXN2* that were identified through literature review and using the gnomAD short tandem repeat (STR) data as a secondary proxy control cohort. We also applied our defined *ATXN2* repeat risk threshold to explore relationships with clinical outcomes.

## Materials and Methods

### Literature review

We reviewed the available peer-reviewed literature to identify all manuscripts presenting analyses assessing *ATXN2* CAG trinucleotide repeat lengths in case-control cohorts of individuals with ALS using the keywords “amyotrophic lateral sclerosis” & “*ATXN2*” or “motor neuron disease” and “*ATXN2*” (as of May 1, 2025). The identified manuscripts were then further refined to those that included statistical association analyses comparing the number of individuals with ALS carrying *ATXN2* repeats of “intermediate” lengths of varying sizes to the number of controls, and odds ratios, confidence intervals, and p-values were extracted, as available. Manuscripts were only considered if p-values for the analyses were presented. Statistical tests applied by each manuscript’s analyses are outlined in Supplemental Table 1. For any manuscript that did not include the odds ratios or confidence intervals, results were only included if carrier counts and total sample sizes were available to manually calculate the values^17^.

We further assessed the peer-reviewed manuscripts to identify those that included the exact carrier counts in individuals with ALS and controls of *ATXN2* repeat expansions for binned lengths ranging from 24 repeats to 34 or more repeats. Manuscripts were still included in this assessment if carriers of a specific repeat length were not identified (e.g. if carrier counts were presented for individuals with ALS and controls for repeat lengths of 24, 25, 26, 27, 28, 29, 30, 33, and 34, but not for repeat lengths of 31 and 33), as it was assumed there were no carriers of the missing repeat lengths within the range of 24 to ≥34 repeats. Cohorts from manuscripts were excluded if there was deemed a potential for sample overlap with another manuscript’s cohort, with the larger cohort retained for analyses.

### ATXN2 repeat lengths in Project MinE

The Project MinE ALS Consortium dataset consists of whole genome sequencing of 6439 individuals with ALS and 2470 unrelated controls. Full details of the sequencing methodology and quality control were previously described^16,18^, and all participants provided written informed consent with study approval from the institutional review board of the University Medical Center Utrecht. Principal component analysis was used to capture variance in population structure of the cohort, and all samples were assessed for pathogenic variants in known ALS-associated genes such as *ALS2*, *ANG*, *C9orf72* (hexanucleotide repeat expansion), *FUS*, *NEK1*, *SOD1*, *TARDBP*, and *VAPB*^18^.

Previously, ExpansionHunter (v5.0)^19^ was used to screen for STR expansions within 1 Mb of the top ALS-associated single nucleotide polymorphisms, using STRs from 18 high-quality genomes and the GangSTR catalog^20^, excluding homopolymers. STR sizes were recoded into biallelic variants using a sliding-window approach. Following quality control of the STR expansion calls, as previously described^21^, 6210 individuals with ALS and 2400 controls remained for analysis. *ATXN2* CAG repeat expansion sizes were extracted from the STR dataset.

Carrier counts were derived for each repeat length from 24 to 34 and for cumulative bins of repeat lengths defined by the lower limit of repeats included in that bin from ≥24 repeats to ≥34 repeats for the individuals with ALS and control cohorts. To quantify any enrichment of *ATXN2* repeat lengths in individuals with ALS compared to controls, we applied logistic regression using Firth’s penalized likelihood for each of the single repeat length from 24 to 34 and for the cumulative repeat length bins from ≥24 to ≥34. To prioritize capturing the true impact of *ATXN2* repeat expansions, individuals with ALS and controls that carried known ALS-associated pathogenic variants in the Project MinE ALS Consortium cohort were excluded from this ALS risk analysis and the subsequent ALS-risk meta-analyses. Models were adjusted for sex and the first three principal components (PCs) to control for population structure, and maximum iterations were increased to ensure convergence for all models (maximum iterations = 1000, maximum step size = 0.2). Odds ratios and 95% confidence intervals were derived from penalized regression coefficients. To account for multiple testing, p-values were adjusted using the Benjamini–Hochberg false discovery rate (FDR) procedure. Significance was measured at an alpha-level of 0.05.

### ATXN2 repeat lengths meta-analyses

Using the 19 peer-reviewed manuscripts that included carrier counts of *ATXN2* repeat lengths in cohorts of individuals with ALS and controls and the Project MinE ALS Consortium cohort, a meta-analysis was conducted. Relative risk for *ATXN2* repeat carrier status and 95% confidence intervals were calculated. The effect estimates were calculated as relative risks, as the meta-analysis was based on aggregated carrier counts comparing *ATXN2* repeat allele frequencies between the various cohorts of individuals with ALS and controls. In contrast, the previous analysis of the Project MinE cohort produced odds ratios that were estimated using individual-level regression-based models adjusting for covariates. Where any cell count was 0, a continuity correction of 0.5 was added to all four cells. The Wald Z-test was applied to each study to test the significance of associations. Pooled estimates were generated with both fixed and random effects models for the meta-analysis, and between study heterogeneity was assessed using the I^2 statistic and Cochran’s Q test. Although some cohorts had low heterogeneity, the presence of studies with significant variation (Q-test p < 0.05) suggested the use of random effects models were most appropriate for the purposes of the meta-analyses.

Meta-analyses were performed separately for each *ATXN2* repeat length from 24 to ≥34 repeats using random effects models. If any of the 20 cohorts did not contain either individuals with ALS or controls carrying *ATXN2* repeats of a specific length, the cohort was excluded from that specific meta-analysis. P-values were adjusted using the Benjamini–Hochberg FDR procedure. Significance was measured at an alpha-level of 0.05.

A secondary analysis of the 20 meta-analysed cohorts was also applied using the gnomAD v4.1.0 STR data as a secondary proxy control cohort^22^. The STR genotype data for *ATXN2* were extracted from the dataset retrieved from gnomAD in May 2025. Data were filtered to only include those with a passing quality filter. Repeat lengths of the individual alleles were extracted from the genotypes, resulting in a dataset of 32,650 total alleles from 16,325 samples.

Pooled relative risk and 95% confidence intervals were calculated to compare the carrier counts for each *ATXN2* repeat length from 30 to ≥34 repeats in the individuals with ALS from the 20 cohorts (n = 20,010) to the gnomAD cohort (n = 16,325). The Wald Z-test was applied to test the significance of associations, p-values were adjusted using the Benjamini–Hochberg FDR procedure, and significance was measured at an alpha-level of 0.05. These comparisons were not part of the weighted meta-analysis but served as an external benchmark, allowing us to evaluate whether the observed enrichments in individuals with ALS held when compared to a general population dataset.

### ALS clinical outcomes

Finally, we assessed relationships between the *ATXN2* CAG trinucleotide repeat expansion lengths in individuals with ALS and key clinical outcomes, including age of onset, survival duration, and diagnostic delay, using the Project MinE ALS Consortium dataset. Analyses were performed on an individual level and all individuals with ALS with *ATXN2* repeat lengths defined for both alleles and data for the relevant clinical outcome measure were included in the analyses. For each clinical parameter, we performed two analyses: 1) a binary analysis comparing the clinical outcomes of those with *ATXN2* repeat lengths of ≥30 to those with lengths <30 and 2) a continuous analysis considering each individual’s maximum raw *ATXN2* repeat length (e.g. the maximum raw *ATXN2* repeat length an individual with repeat lengths of 31 and 26 was considered 31).

Linear regressions were used to model age at ALS symptom onset as a function of *ATXN2* repeat length, adjusting for sex, and carrier status of other known ALS-associated pathogenic variants. Due to the known association between family history of ALS and age of ALS onset, individuals with ALS with a first-degree family history of the disease were excluded from the age of symptom onset analyses. As described above, individual models were used to capture *ATXN2* repeat lengths as a binary and as a continuous variable.

Cox proportional hazard models were used to assess associations between *ATXN2* repeat length and survival period in years from symptom onset to death or last follow-up, adjusting for age at onset, sex, and carrier status of other known ALS-associated pathogenic variants. As the proportional hazards assumption was violated for age at onset when modeled as a continuous variable (p = 3.2e-07), the model was stratified by appropriately distributed age at onset bins (<50 years, 50-60 years, and >60 years), such that the baseline hazard would capture the variance by age at onset without estimating specific hazard ratios. Kaplan–Meier survival curves were plotted for individuals with *ATXN2* repeat lengths <30 and ≥30 and were annotated with p-values derived from the Cox model.

Distributional assumptions were evaluated for diagnostic delay and identified a substantial positive skew (skewness = 7.08) and non-normality based on the Anderson-Darling test (A = 509.55, p < 2.2e-16). Multiple linear regression models were used to identify the model of best fit to assess associations between *ATXN2* repeat length and diagnostic delay in years, adjusting for age at onset, sex, carrier status of other known ALS-associated pathogenic variants, and site of onset (using spinal onset as a reference). We compared a standard linear regression using the raw diagnostic delay values, a linear regression using the log-transformed delay values, and a generalized linear model with Gamma distribution and log link. Model fit was assessed using the Akaike Information Criterion (AIC). The log-transformed linear model yielded the lowest AIC (AIC = 11719.90), compared with the raw linear model (AIC = 77399.01) and the gamma model (AIC = 71031.70). Based on these results, the log-transformed modeling of diagnostic delay was applied. Again, separate models were applied capturing *ATXN2* repeat lengths as a binary and a continuous variable.

### Statistical analyses and data visualization

Statistical analyses were performed using the R statistical software v4.2.1^23^ in R Studio v1.4.17. Meta-analyses were performed using the *metafor* R package (v4.8.0)^24^. Cox proportional hazard modeling was performed using the *survival* R package (v3.3.1)^25^ and results were visualized using the *survminer* R package (v0.5.0)^26^. All other data visualizations were performed using the *ggplot2* R package (v3.4.4)^27^.

## Results

### Review of the current ALS ATXN2 literature

To assess the current understanding of the ALS risk imparted by varying *ATXN2* CAG trinucleotide repeat expansion lengths, we surveyed the literature for all manuscripts presenting an analysis of *ATXN2* repeat lengths in case-control cohorts of individuals with ALS. We identified 34 manuscripts published between 2010 to 2025, 30 of which included statistical analyses comparing the number of individuals with ALS carrying *ATXN2* of “intermediate” repeat lengths of varying sizes to the number of controls^2–15,28–43^. We extracted the odds ratios, confidence intervals, and p-values from the identified manuscripts to compare the associations between *ATXN2* repeats of various lengths and ALS (Supplemental Figure 1; Supplemental Table 1).

### ATXN2 repeats in the Project MinE ALS Consortium dataset

Following quality control of the Project MinE ALS Consortium ExpansionHunter analysis, *ATXN2* CAG trinucleotide repeat expansion lengths were defined in 6210 individuals with ALS and 2400 controls (Figure 1). Demographics of the cohort are presented in Table 1.

**Figure 1.**
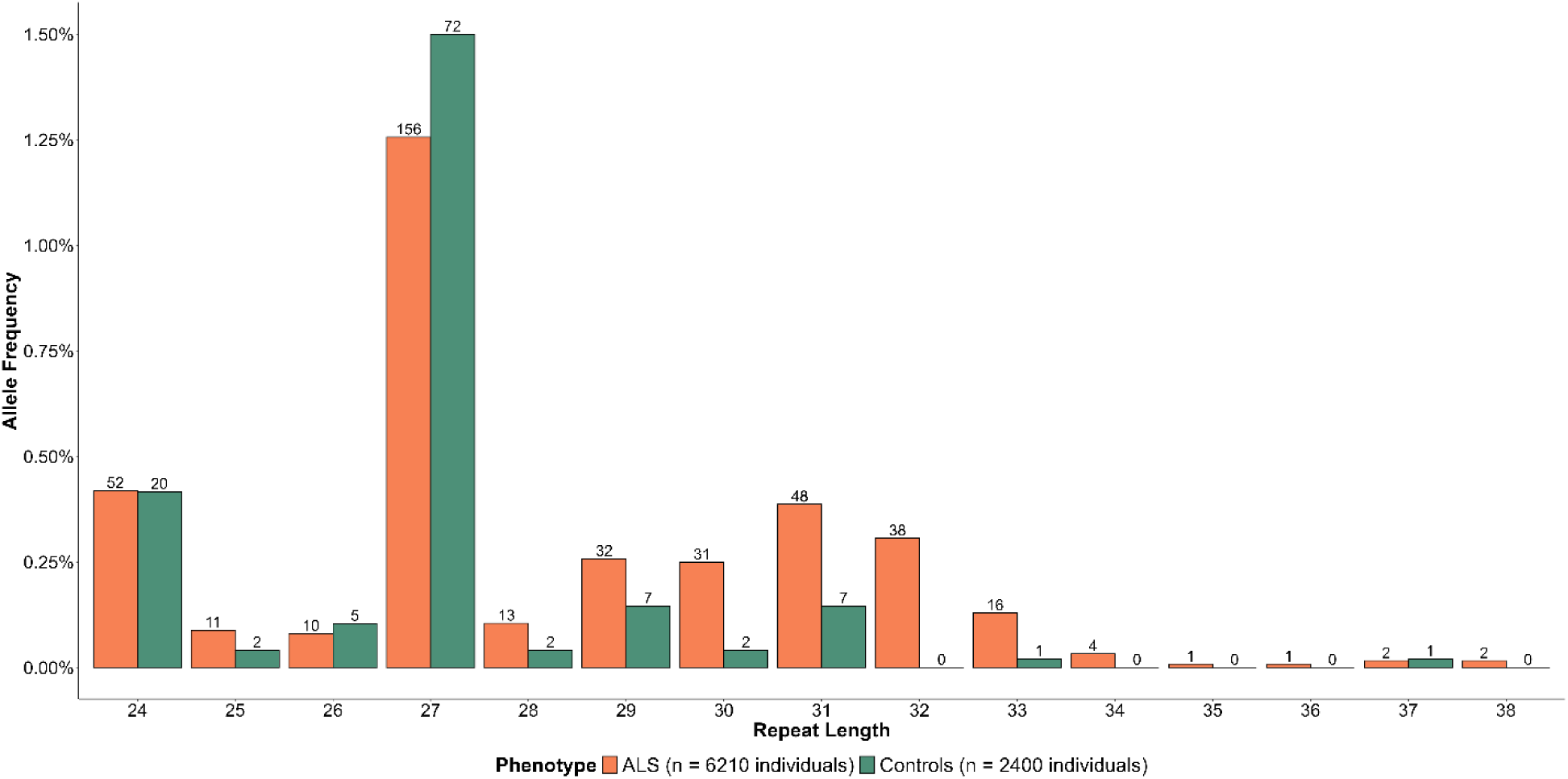
Distribution of *ATXN2* alleles with repeat lengths of 24 or more in the Project MinE Sequencing Consortium dataset (individuals with ALS = 6210; controls = 2400). There were 12003 alleles of size 23 or fewer repeats in individuals with ALS and 4681 alleles of size 23 or fewer repeats in controls.

**Table 1.** Demographics of the 6210 individuals with ALS and 2400 controls encompassed by The Project MinE ALS Consortium that had *ATXN2* CAG trinucleotide repeat expansion lengths successfully defined using ExpansionHunter analysis.

|  | Individuals with<br>ALS | Controls |
| --- | --- | --- |
| **Total n** | 6210 | 2400 |
| **Sex at birth** |  |  |
| Female | 2496 (40.2%) | 1131 (47.1%) |
| Male | 3714 (59.8%) | 1269 (52.9%) |
| **Cohort** |  |  |
| Belgium | 534 (8.6%) | 178 (7.4%) |
| France | 232 (3.7%) | 36 (1.5%) |
| Ireland | 462 (7.4%) | 231 (9.6%) |
| Israel | 100 (1.6%) | 0 |
| Italy | 55 (0.9%) | 0 |
| Netherlands | 1780 (28.7%) | 1040 (43.3%) |
| Portugal | 47 (0.8%) | 13 (0.5%) |
| Spain | 358 (5.8%) | 154 (6.4%) |
| Sweden | 203 (3.3%) | 111 (4.6%) |
| Switzerland | 44 (0.7%) | 0 |
| Turkey | 601 (9.7%) | 132 (5.5%) |
| United Kingdom | 1376 (22.2%) | 439 (18.3%) |
| United States | 418 (6.7%) | 66 (2.7%) |
| **Carriers of ALS-associated pathogenic variants<sup>a</sup>** | 388 (6.2%) | 9 (3.7%) |
| **First-degree family history of ALS** |  |  |
| Yes | 189 (3.0%) | 2 (0.1%) |
| Unknown | 3391 (54.6%) | 2240 (93.3%) |
| **Age at symptom onset (mean ± sd)** | 60.3 ± 12.6 years | - |
| **Age at diagnosis (mean ± sd)** | 62.6 ± 11.8 years | - |
| **Diagnostic delay (mean ± sd)** | 455.6 ± 520.8 days | - |
| **Site of onset** |  |  |
| Bulbar | 1658 (26.7%) | - |
| Generalized | 210 (3.4%) | - |
| Spinal | 4062 (65.4%) | - |
| Thoracic/respiratory | 107 (1.7%) | - |
| Unknown | 173 (2.8%) | - |
| **Survival time/time to last follow-up (mean ± sd)** | 3.6 ± 3.5 years | - |
| **Survival status (by last follow-up)** |  |  |
| Deceased | 4541 (73.1%) | - |
| Alive | 1511 (24.3%) | - |
| Unknown | 158 (2.5%) | - |

<sup>a</sup>Pathogenic variants were defined as those in known ALS-associated genes such as *ALS2*, *ANG*, *C9orf72* (hexanucleotide repeat expansion), *FUS*, *NEK1*, *SOD1*, *TARDBP*, and *VAPB* and were defined in previous analyses.

To analyze the contribution of various repeat lengths to the risk of ALS, we applied allele-based Firth’s penalized likelihood models, adjusting for sex and PCs 1-3. Individuals with ALS and controls known to carry ALS-associated pathogenic variants were excluded for these ALS risk models (individuals with ALS = 5822, controls = 2391). We first used individual models for each repeat length from 24 repeats to 34 repeats, comparing the number of alleles observed in individuals with ALS to controls (Figure 2). We found that *ATXN2* repeats of lengths 30 (OR = 4.72 [1.56-23.23], p = 3.62e-3, FDR-p = 1.99e-2), 31 (OR = 2.49 [1.22-5.89], p = 1.05e-2, FDR-p = 3.86e-2), and 32 (OR = 32.61 [4.64-4125.62], p = 1.77e-6, FDR-p = 1.94e-5) were all significantly enriched in individuals with ALS compared to controls following multiple test corrections.

**Figure 2.**
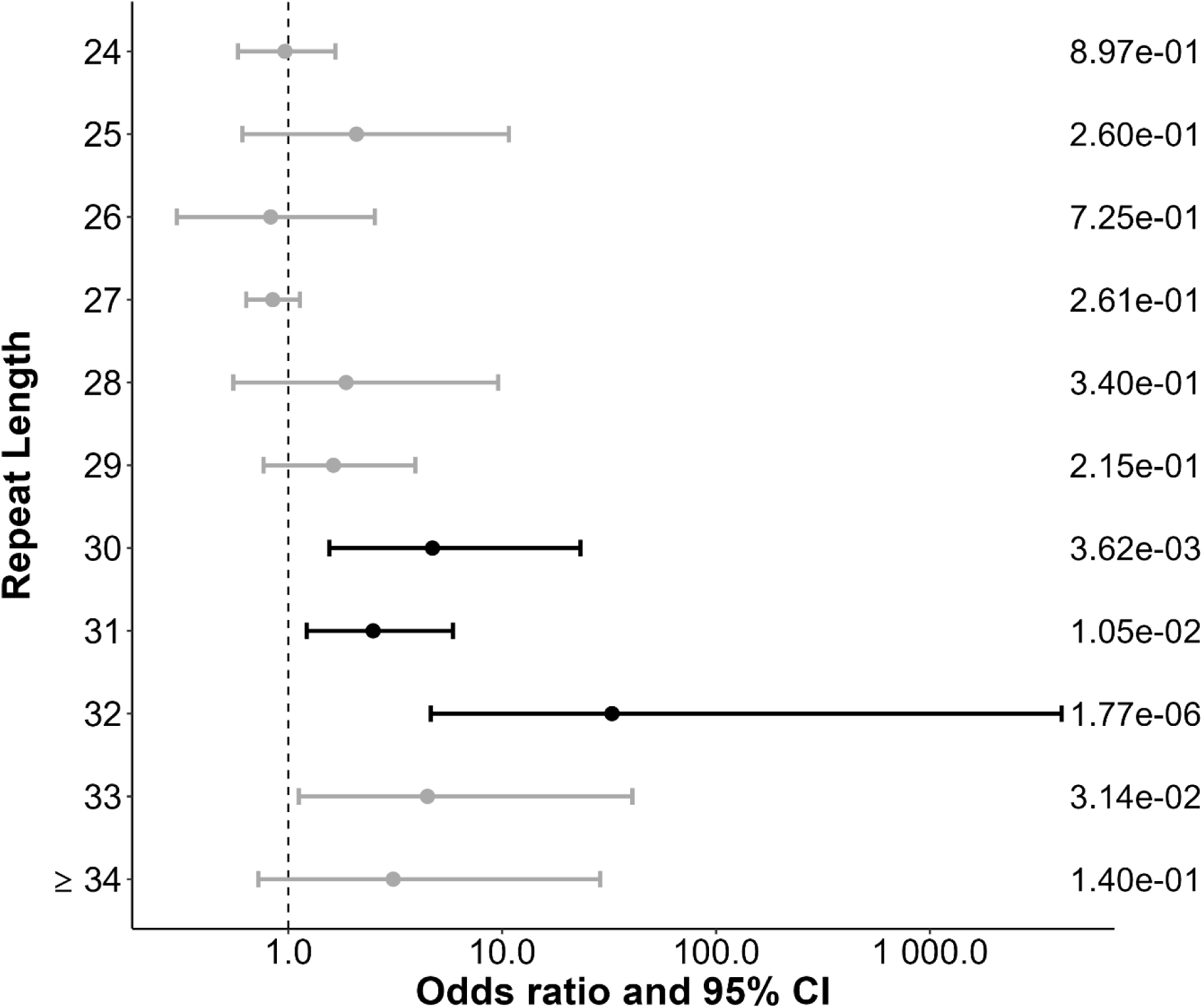
Enrichment analysis of *ATXN2* alleles with repeat lengths of 24 or more in the Project MinE Sequencing Consortium dataset (individuals with ALS = 5822; controls = 2391). Firth’s penalized likelihood models, adjusted for sex and PCs 1-3 were used to compare the number of alleles of each individual *ATXN2* repeat length. Raw p-values are displayed on the right side of the plot. Odds ratio and 95% confidence intervals (CIs) displayed in black indicate significance at an alpha level of 0.05 following the Benjamini–Hochberg false discovery rate (FDR) procedure.

We also assessed the *ATXN2* repeat lengths using bins defined by the lower limit of repeats included in that bin from ≥24 repeats to ≥34 repeats (e.g. ≥24 refers to alleles with lengths of 24 repeats or more). Here, all repeat bins were significantly enriched in individuals with ALS following multiple test corrections, aside from alleles of ≥34 repeats (Supplemental Figure 2). However, we propose that these results were likely due to the increased risk of ALS imparted by the higher repeat sizes, specifically, ≥30 repeats based on analysis of the individual repeat lengths, rather than true associations of ALS risk imparted from *ATXN2* repeats of lengths ≥24.

### Meta-analysis of ATXN2 repeat lengths in ALS

We next surveyed the 34 manuscripts described above within the peer-reviewed literature that included *ATXN2* repeat lengths of cohorts of individuals with ALS and controls and found 19 that included the allele counts of repeat lengths from 24 repeats to ≥34 repeats and did not seemingly include overlapping individuals. Using these 19 cohorts, we meta-analyzed the allele counts of individual *ATXN2* repeat expansions ranging from lengths 24 to ≥34 with the data from the Project MinE ALS Consortium, resulting in a total analysis of 19212 individuals with ALS in comparison to 22177 controls (Supplemental Figure 3; Supplemental Table 2).

In alignment with the results we observed using the Project MinE ALS Consortium dataset individually, the meta-analysis applying random effects models revealed that, indeed, *ATXN2* repeat expansions of lengths 30 (RR = 2.48 [1.31-4.69], p = 5.14e-3, FDR-p = 1.13e-02), 31 (RR = 3.29 [2.12-5.10], p = 9.74e-8, FDR-p = 5.36e-07), 32 (RR = 8.51 [4.78-15.15], p = 3.47e-13, FDR-p = 3.81e-12), 33 (RR = 6.13 [3.04-12.34], p = 3.74e-7, FDR-p = 1.37e-06), and ≥34 (RR = 4.09 [1.90-8.81], p = 3.10e-4, FDR-p = 8.52e-04) were significantly enriched in individuals with ALS in comparison to controls (Figure 3).

**Figure 3.**
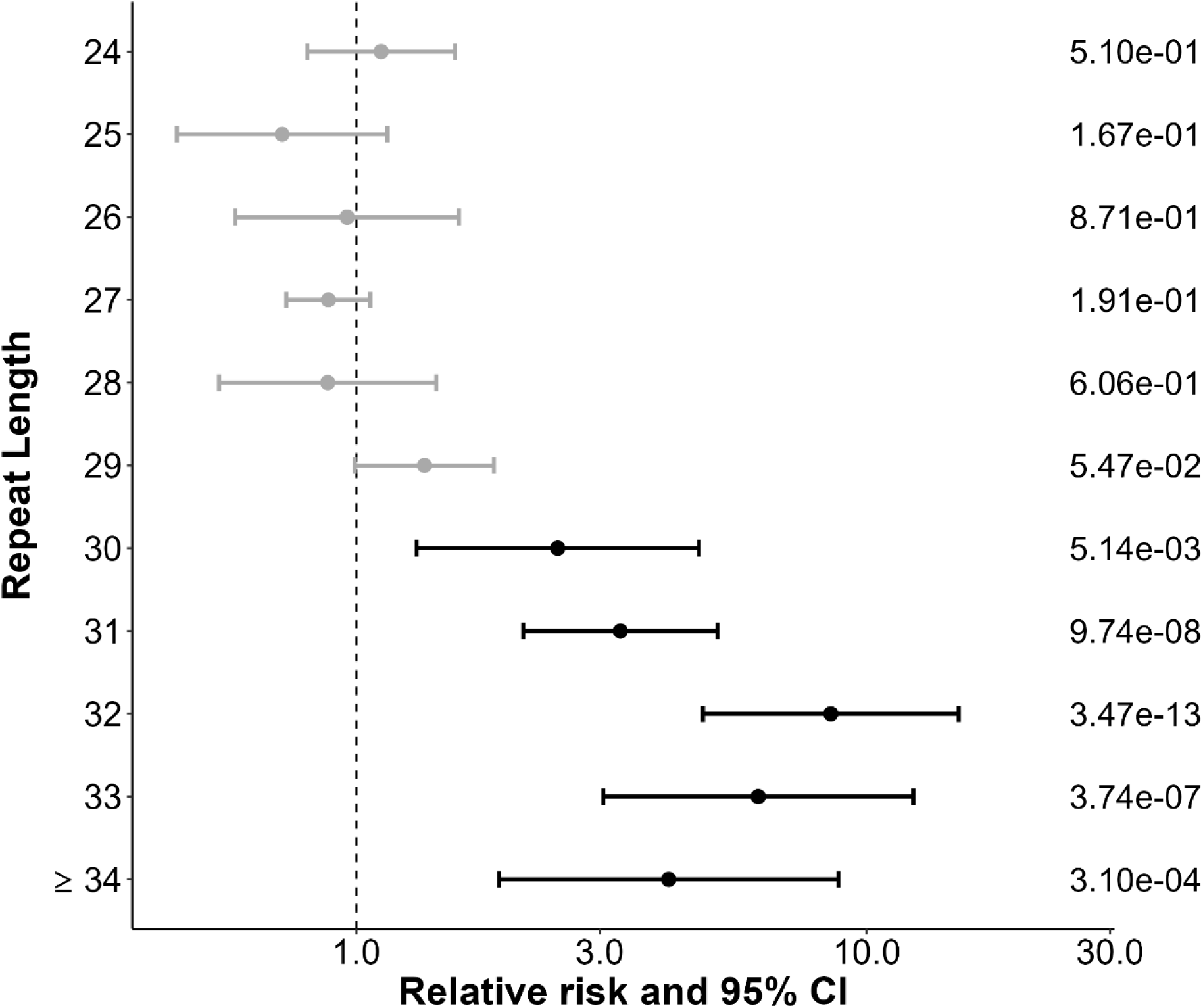
Isolated meta-analysis results of 19 studies presenting *ATXN2* repeat lengths of individuals with ALS and controls with the Project MinE Sequencing Consortium Dataset (individuals with ALS = 19,212; controls = 22,177). Relative risk and 95% confidence intervals were calculated for each of the 20 total cohorts, applying a continuity correction (0.5 to each cell) when counts were zero. The Wald Z-test was applied to each study to test the significance of associations. Pooled estimates were generated using a random effects model following assessment using the the I^2 statistic and Cochran’s Q test. Studies were excluded from the analysis of a specific repeat length if there were no individuals with ALS or controls with repeats of that specific length. Raw p-values are displayed on the right side of the plot. Odds ratio and 95% confidence intervals (CIs) displayed in black indicate significance at an alpha level of 0.05 following the Benjamini–Hochberg false discovery rate (FDR) procedure.

### Secondary analysis of ATXN2 repeat lengths using gnomAD

Finally, we used the gnomAD v4.1.0 STR data as a secondary proxy control cohort to compare the frequency of *ATXN2* repeat lengths to the individuals with ALS encompassed by the 20 meta-analyzed cohorts to strengthen our evidence suggesting repeats ≥30 are associated with ALS risk (Figure 4A). The pooled relative risk and 95% confidence intervals were calculated to compare the alleles carried by individuals with ALS from the 20 cohorts (n = 19212) to the gnomAD cohort (n = 16325) and Wald Z-tests were applied to determine the significance of associations (Figure 4B). Again, we confirmed that individuals with ALS were enriched for *ATXN2* repeat expansions of lengths ≥30 in comparison to the gnomAD proxy control cohort.

**Figure 4.**
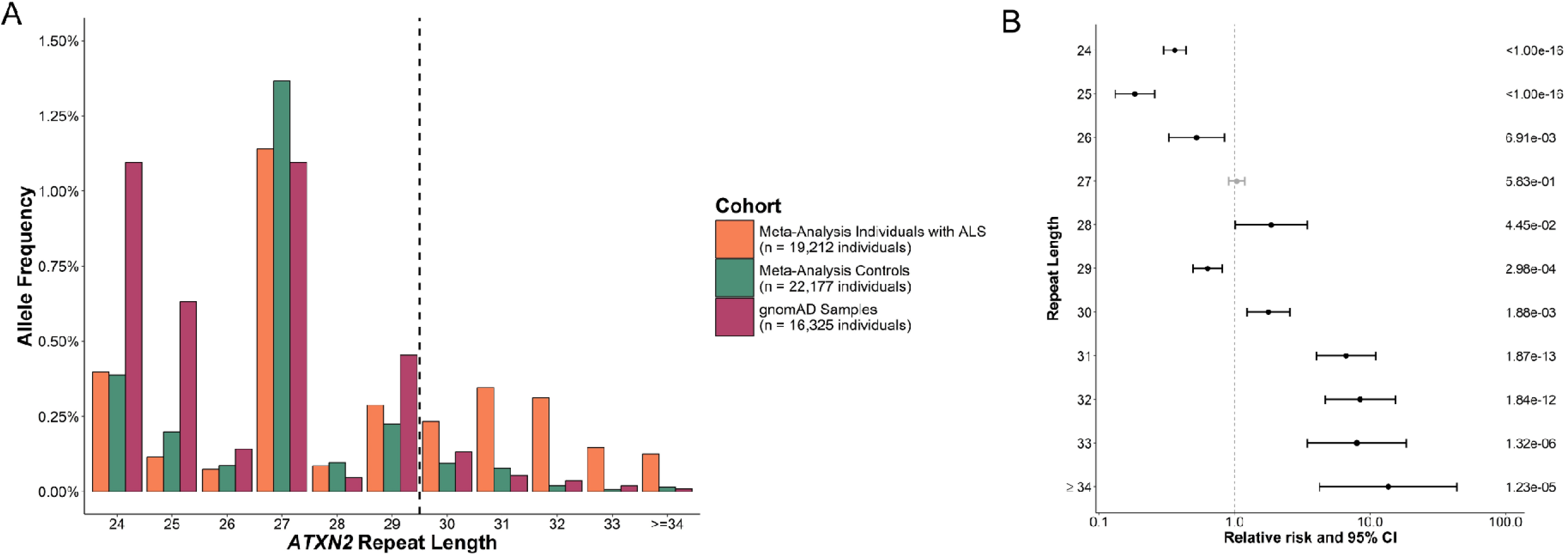
Secondary analysis of the *ATXN2* repeat lengths from the 20 studies included in the meta-analysis using the gnomAD v4.1.0 tandem repeat dataset (n = 16,325) as a proxy control cohort. **A)** Distribution of *ATXN2* alleles with repeat lengths of 24 or more in the total meta-analysis cohort and the gnomAD tandem repeat dataset. The dashed line represents the lower threshold of repeats that suggest significantly increased risk of ALS based on the meta-analysis results. **B)** Enrichment analysis of *ATXN2* alleles with repeat lengths of 30 or more in the pooled individuals with ALS from the meta-analysis (n = 19,212) and the gnomAD v4.1.0 tandem repeat dataset as a proxy control cohort (n = 16325). The pooled relative risk and 95% confidence intervals were calculated, and the Wald Z-test was applied to test the significance of associations. Raw p-values are displayed on the right side of the plot. Odds ratio and 95% confidence intervals (CIs) displayed in black indicate significance at an alpha level of 0.05 following the Benjamini–Hochberg false discovery rate (FDR) procedure.

### Contribution of ATXN2 repeat expansions to clinical outcomes

Now that a threshold of ≥30 *ATXN2* repeats was established as significantly contributing to ALS risk, we sought to determine whether the risk factor was also associated with clinical parameters. For each clinical parameter, we performed two individual-level analyses: 1) a binary analysis comparing the clinical outcomes of those with *ATXN2* repeat lengths of ≥30 to those with lengths <30 and 2) a continuous analysis considering each individual’s maximum raw *ATXN2* repeat length.

We first applied a linear regression to assess the contribution of *ATXN2* repeat length on age of ALS onset, adjusting for sex and the presence of other known ALS-associated pathogenic variants (n = 5853 individuals with ALS; Figure 5A). In both the binary and continuous analysis, *ATXN2* repeat lengths were not found to significantly contribute to the age of ALS onset (binary *ATXN2* variable, β = 1.82 [−0.34-3.99], p = 9.86e-2; continuous *ATNX2* variable, β = 0.10 [−0.08-0.29], p = 2.78e-1).

**Figure 5.**
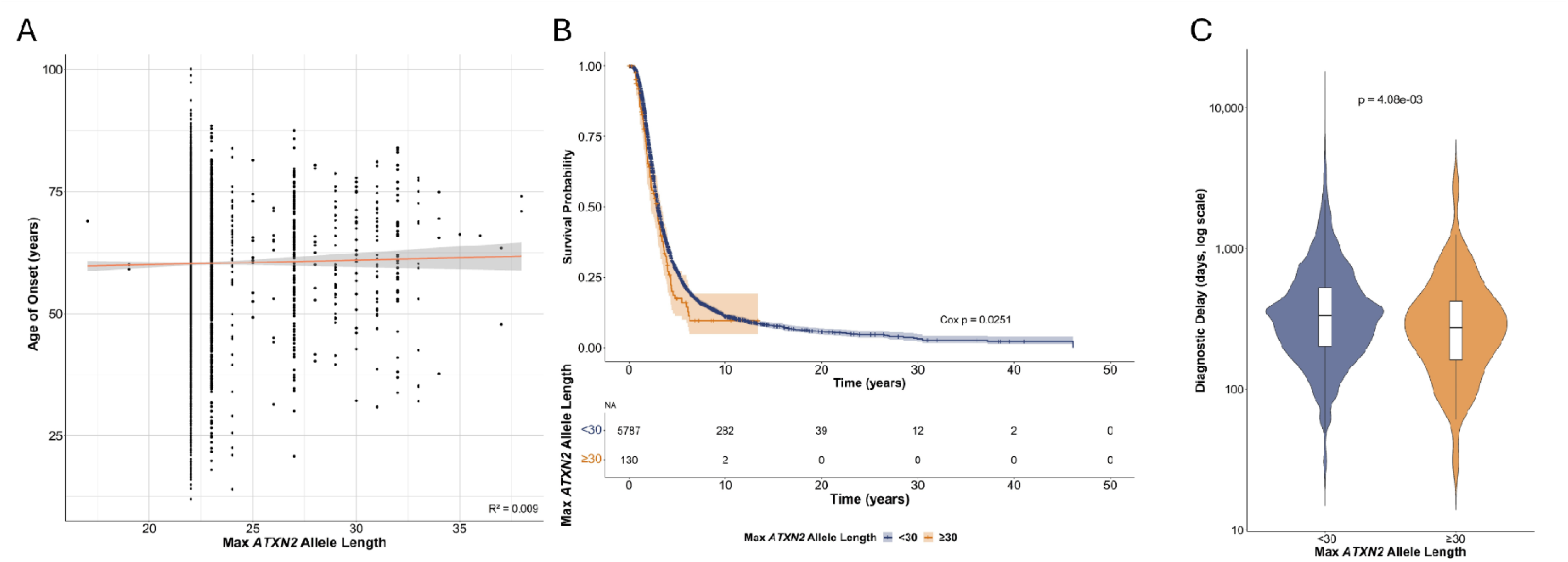
Influence of *ATXN2* expansion repeat length on the clinical parameters of individuals with ALS in the Project MinE ALS Consortium dataset. A) Age of onset in individuals with ALS (n = 5853) based on their maximum *ATXN2* allele repeat expansion length. R^2^ represents the unadjusted correlation between the raw maximum *ATXN2* repeat lengths and ages of onset, which was not significant in a linear regression model following adjustment for sex and the presence of other known ALS-associated pathogenic variants. Individuals with a first-degree family history of ALS were excluded from this analysis. B) Kaplan-Meier survival curves stratified by maximum *ATXN2* allele repeat length. Survival probability over time is showed for individuals with *ATXN2* repeat lengths <30 and ≥30, as determined to be the threshold defining ALS risk. Shaded areas represent 95% confidence intervals. The table displays the number of individuals still surviving at each time interval (years). A Cox proportional hazard model adjusting for sex, age of onset (stratified), and the presence of other known ALS-associated pathogenic variants showed a significant inverse association between individuals with ALS’s (n = 4439) maximum raw *ATXN2* repeat length and survival period (HR = 1.02 [1.00-1.04], p = 2.51e-2). C) Violin plot of individuals with ALS’s diagnostic delay (years) compared to the maximum length of their *ATXN2* repeat expansion binned based on our predetermined risk threshold. A log-transformed linear regression model found that individuals with ALS (n = 5046) with a maximum *ATXN2* allele ≥30 repeats in length had significantly shorter diagnostic delays than individuals with <30 repeats (β = −0.21 [−0.37-(−0.07)], p = 4.08e-3).

We next applied a Cox proportional hazard model to assess the contribution of *ATXN2* repeat length on survival period, adjusting for sex, age of onset (stratified), and the presence of other known ALS-associated pathogenic variants (n = 4439 individuals with ALS; Figure 5B). Here, we did not observe a significant relationship between the binary *ATXN2* variable and survival period, such that individuals with ALS with *ATXN2* repeat lengths of ≥30 did not display significantly different survival periods to those with lengths <30 (HR = 1.22 [0.99-1.50], p = 5.86e-2); although, individuals with ALS with an expansion of ≥30 repeats displayed a mean survival period of 2.76 years, while those with an expansion of <30 repeats displayed a mean survival period of 3.65 years. When the contribution of the maximum raw *ATXN2* repeat length to survival period was assessed as a continuous variable, we found that repeat length was significantly inversely associated with survival period (HR = 1.02 [1.00-1.04], p = 2.51e-2).

Finally, due to the extreme non-parametric bias observed for diagnostic delay, we examined several linear regression models and assessed their AIC to define a log transformed regression as the model of best fit to assess associations with this clinical outcome, adjusting for sex, age of onset, site of onset, and the presence of other known ALS-associated pathogenic variants (n = 5046 individuals with ALS; Figure 5C). We observed that individuals with ALS with *ATXN2* repeat lengths of ≥30 experienced significantly shorter diagnostic delays compared to those with lengths <30 (β = −0.21 [−0.37-(−0.07)], p = 4.08e-3), with average diagnostic periods of 375.41 days and 457.34 days, respectively. Similarly, raw maximum *ATXN2* repeat lengths were significantly inversely associated with diagnostic delay (β = −0.01 [−0.03-(−8.5e-4)], p = 3.65e-2).

## Discussion

Since the first association made in 2010, there has been great debate regarding the number of *ATXN2* CAG trinucleotide expansion repeats that confer ALS risk, ranging from a lower limit of 27 to 31 repeats ^7,14,44^. Yet the lack of consensus presents challenges within the field, particularly to meet the need of rapidly expanding clinical genetic testing efforts for individuals with ALS and guidance in inclusion criteria of any future targeted therapeutic strategies. Here, we present the largest meta-analysis of *ATXN2* repeat lengths to date, encompassing 19212 individuals with ALS and 22177 controls, defining risk from *ATXN2* trinucleotide expanded alleles in ALS as those ≥30 repeats in length.

We identified 19 studies that presented the carrier counts of expansions ranging from 24 to ≥34 repeats in cohorts of individuals with ALS and controls that we meta-analysed with the *ATXN2* repeat lengths of the large-scale Project MinE ALS Consortium dataset. Interestingly, our amalgamation of large-scale efforts across the ALS genomics literature has presented a lower limit of 30 repeats defining ALS risk, representing a compromise between two of the most commonly referenced studies found within the current peer reviewed literature, which suggested lower limits of 29 repeats and 31 repeats ^7,14^. This finding was also consistent with the prior analysis of the Project MinE Consortium dataset by van Vugt *et al*. who identified ≥30 repeats as the optimal threshold associated with ALS using a best-threshold analysis^21^. Although both studies leveraged the Project MinE cohort, the statistical frameworks of our studies differed, with van Vugt *et al*. evaluating optimal thresholds of binned repeat lengths and our analysis assessing individual repeat lengths. Importantly, the Project MinE cohort represented one of 20 independent cohorts included in the random-effects meta-analysis, which allowed for the integration of evidence across multiple populations and studies despite differences in cohort characteristics, such as sample size and case-control balance. To further validate the associations observed beginning at 30 repeats in length, we also performed a secondary assessment of the individuals with ALS captured within the meta-analysis using gnomAD as a proxy control cohort, which again demonstrated significant enrichments of all *ATXN2* repeat lengths from 30 to ≥34 repeats in individuals with ALS.

In alignment with the previous largest meta-analysis presented by Spoviero *et al*. we found that risk of ALS increased with allele repeat size until a length of 33 repeats, where a drop off in risk was observed, likely attributed to individuals with 33 or more repeats that would instead develop spinocerebellar ataxia ^7,45^. Yet, the enrichment of *ATXN2* expansions of ≥34 repeats in individuals with ALS compared to controls remained significant even though these lengths are known pathogenic causes of spinocerebellar ataxia. Although uncommon, there have been multiple previous reports of families with members diagnosed with ALS that carry *ATXN2* expansions of ≥34 repeats ^46,47^, as well as a recent analysis that identified full length expansions in individuals with seemingly sporadic ALS^15^. The authors of the latter study concluded that *ATXN2* likely exhibits pleiotropy across neurodegenerative phenotypes, including ALS, spinocerebellar ataxia, and parkinsonism. Based on our results and these previous findings, we only present a lower limit of trinucleotide repeats to be used for defining ALS risk and do not recommend that an upper limit is consider for either research or clinical genetic testing efforts.

We also assessed whether *ATXN2* repeat lengths contributed to key clinical outcome measures in individuals with ALS. As has been previously reported, we did not identify a significant relationship between *ATXN2* repeat length, either as a continuous or binary variable (<30 repeats vs. ≥30 repeats), and age of ALS onset ^7,34,44,48^. However, we did identify a relationship between *ATXN2* repeat lengths as a continuous metric and duration of disease, such that those with longer repeat lengths experienced significantly shorter survival periods. Although this effect remains debated within the literature ^7,13,36^, our results reflected a 2022 analysis by Chio *et al*. that reported a one-year shorter survival period for those carrying an *ATXN2* expansion of ≥31 repeats than those with an expansion <31 repeats ^49^. Similarly, we observed an average survival period of 2.76 years in individuals with ALS with an expansion of ≥30 repeats, while those with an expansion of <30 repeats had an average survival period of 3.65 years. Finally, we found that individuals with ALS with expansions of <30 repeats had significantly greater diagnostic delays than those carrying the risk variant allele of ≥30 repeats; however, we remain cautious in the interpretation of these findings as many confounding factors known to contribute to diagnostic periods could not be considered in the analysis, such as cognitive features, clinical specialty of first assessment, or socio-economic factors ^50,51^.

Although to the best of our knowledge this study represents the largest meta-analysis to date defining a threshold of *ATXN2* repeat lengths that are associated with ALS risk, we recognize limitations remain in the design. We cannot be certain that there were no overlapping participants between the included cohorts. To mitigate this potential, we applied a rigorous review process of the literature, that considered recruitment sites, recruitment periods, overlapping study authorship, and general study methods. When studies had multiple sources of evidence of potential overlap, the largest cohort was chosen for inclusion. Further, we did not account for any effects of CAA interruptions in the CAG trinucleotide repeat expansion sequences, as these were not examined using the ExpansionHunter method applied to the Project MinE ALS Consortium dataset, and the data were unavailable for most cohorts incorporated into the meta-analysis. As the CAA interruptions to the expansions have been found to be potential modifiers of patient phenotypes, including contributing to risk of dopa-responsive parkinsonism and influencing age of ALS onset ^52,53^, the inability to consider their impact in our analyses may have limited our ability to clearly delineate the associations between *ATXN2* and ALS clinical outcomes. We also could not assess the potential contribution of additional genetic risk factors in individuals carrying *ATXN2* repeat expansions, as only two individuals with expansions ≥30 repeats in length also carried a known ALS-associated pathogenic variant. However, oligogenic models of ALS pathogenesis have been previously proposed^54,55^, and future studies with larger cohorts of individuals carrying multiple genetic risk factors will be important to further delineate the involvement of *ATXN2* in these events. Finally, given the variable populations of genetic ancestry in the cohorts included in the meta-analysis, we applied a random-effects model to account for between-study heterogeneity and used the global gnomAD dataset as a secondary proxy control cohort. However, both the cohorts included in the meta-analysis and the global gnomAD dataset display an overrepresentation of European individuals and future analyses investigating the impact of genetic ancestry will be important to refine our understanding of *ATXN2*-associated ALS in diverse populations.

Following years of inconsistent conclusions, our analysis presents a conclusion suggestive of compromise. We propose a lower-limit threshold of ≥30 *ATXN2* trinucleotide repeats in length defines ALS risk as determine by meta-analyses of available ALS case-control cohorts within the literature. This has important implications for clinical genetic testing practices, allowing for improved accuracy in risk interpretation and guidance for patients and their families. More broadly, our results contribute critical confirmation to our understanding of what repeat expansions influence ALS risk to aid in future functional assessments of pathogenic mechanisms, potentially opening avenues for future therapeutic strategies.

## Supporting information

Project MinE ALS Sequencing Consortium

## Data Availability

The data sets used that support the findings in this study are available from The Project MinE consortium public repository. To gain access to the data, an account request must be made to. Data access will require the completion of a data access request. Further information about data access can be found at https://www.projectmine.com/research/data-sharing/. Code used for the analyses can be found at https://github.com/allisondilliott/ATXN2_Meta-Analysis.git.

## Acknowledgements

AAC is an NIHR Senior Investigator (NIHR202421) and a Visiting Professor at the Perron Institute for Neurological and Translational Science, Australia. This is work was partly supported by an EU Joint Programme - Neurodegenerative Disease Research (JPND) project. The project is supported through the UK MND Research Institute, the following funding organisations under the aegis of JPND - www.jpnd.eu *(United Kingdom, Medical Research Council* (MR/L501529/1; MR/R024804/1) *and Economic and Social Research Council* (ES/L008238/1)) and through the Motor Neurone Disease Association, My Name’5 Doddie Foundation, MND Scotland, LifeArc, Alan Davidson Foundation, and Darby Rimmer Foundation. This study represents independent research part funded by the National Institute for Health Research (NIHR) Biomedical Research Centre at South London and Maudsley NHS Foundation Trust and King’s College London.

