## Supplemental for "Assessment of *ATXN2* Repeat Expansion Length and Risk of ALS: A Meta-Analysis"

### Supplemental Material

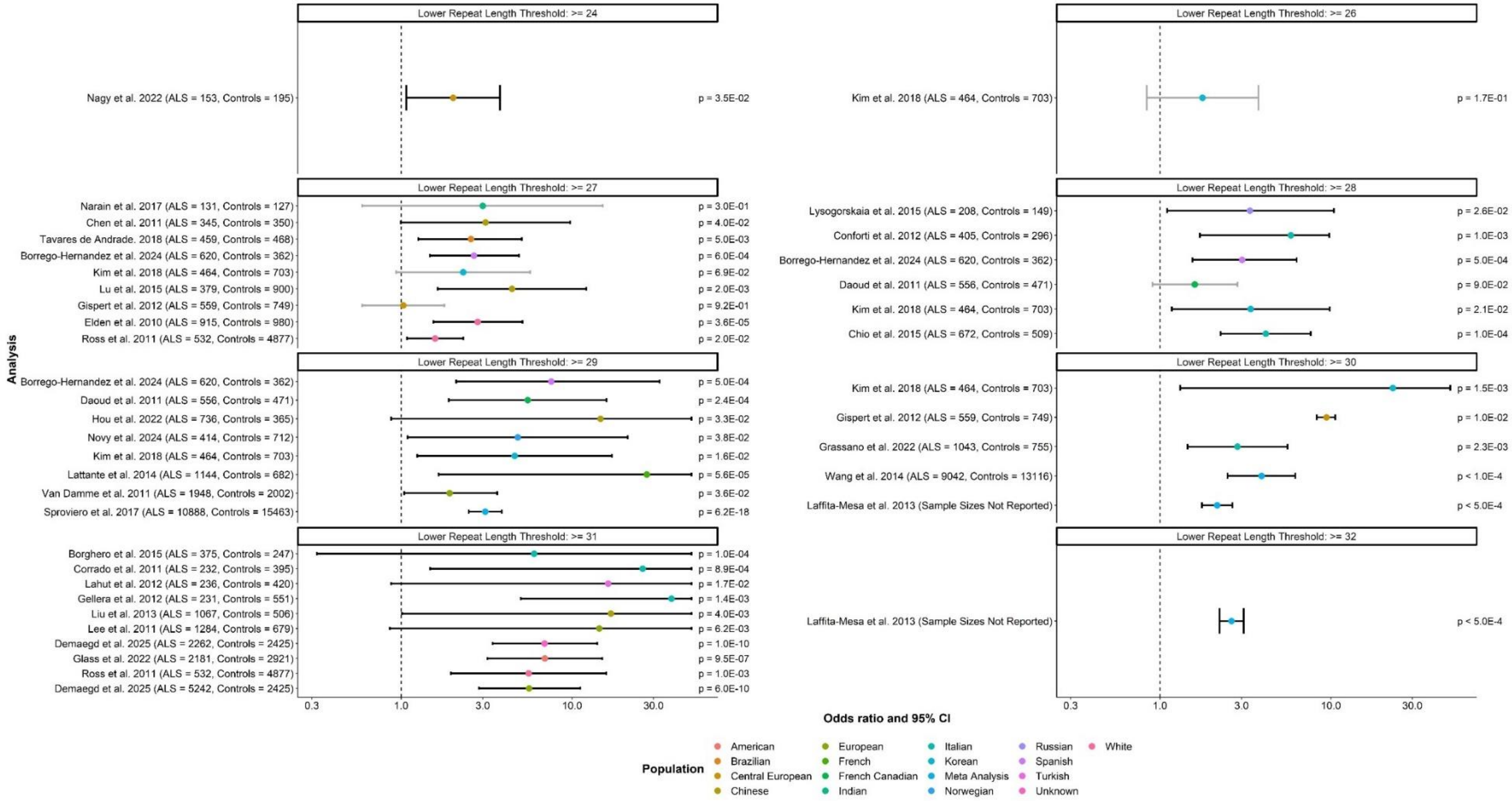


**Supplemental Figure 1. Statistical analyses presented within the currently available peer-reviewed literature presenting associations between *ATXN2* repeat lengths of various sizes and ALS risk.** Of the 34 manuscripts found to present *ATXN2* repeat lengths in case-control cohorts, 30 presented results of statistical association analyses for repeats of at least one length threshold. P-values were pulled directly from the identified manuscripts. Odds ratios and confidence intervals were pulled directly from the identified manuscripts, where available, otherwise were calculated using the presented carrier counts and sample sizes.

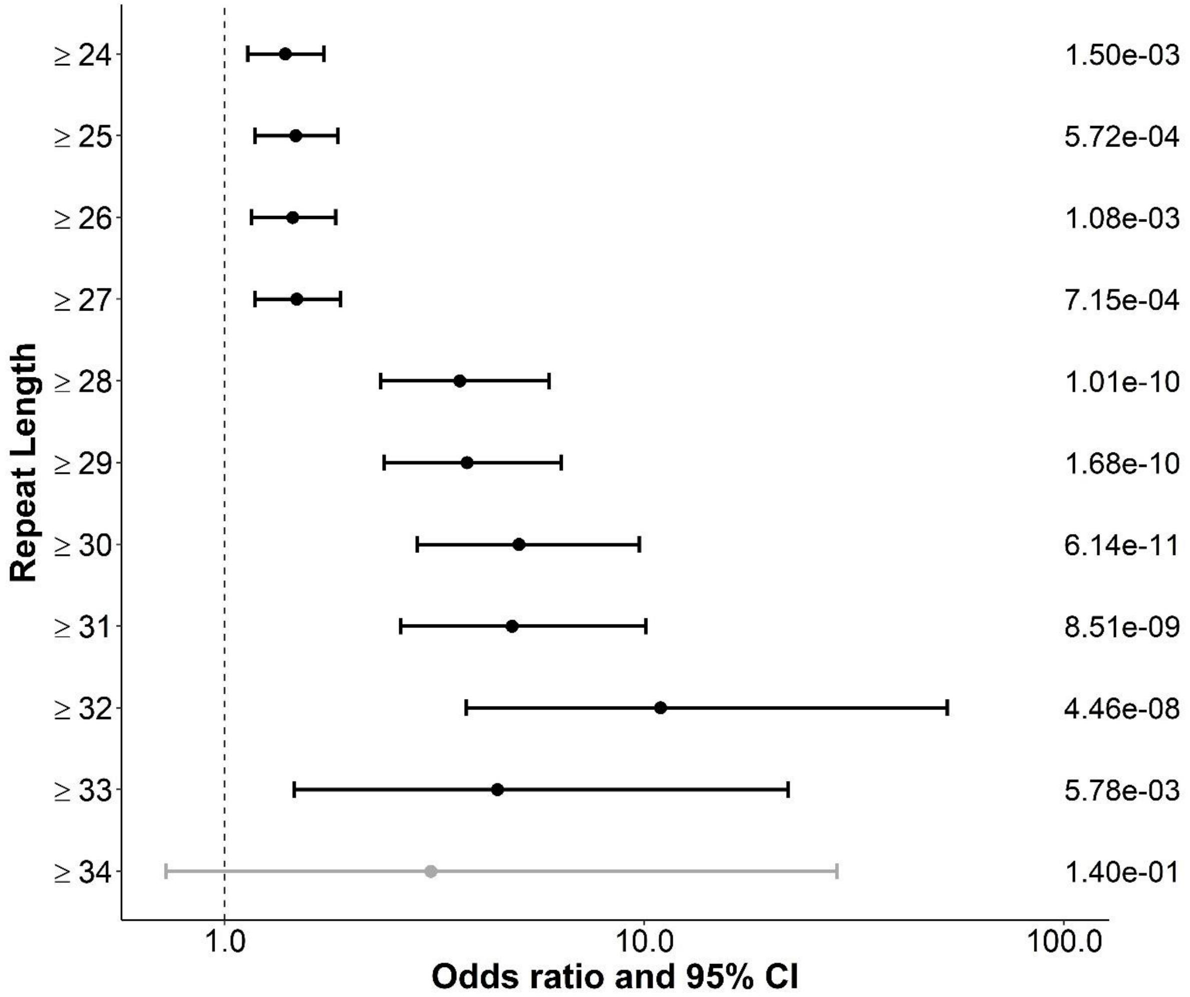


**Supplemental Figure 2. Enrichment analysis of *ATXN2* alleles with repeat lengths of 24 or more in the Project MinE Sequencing Consortium dataset (individuals with ALS = 5822; controls = 2391).** Firth's penalized likelihood models, adjusted for sex and PCs 1-3 were used to compare the number of alleles of binned *ATXN2* repeat length based on a lower limit size threshold. Raw p-values are displayed on the right side of the plot. Odds ratio and 95% confidence intervals (CIs) displayed in black indicate significance at an alpha level of 0.05 following the Benjamini–Hochberg false discovery rate (FDR) procedure.

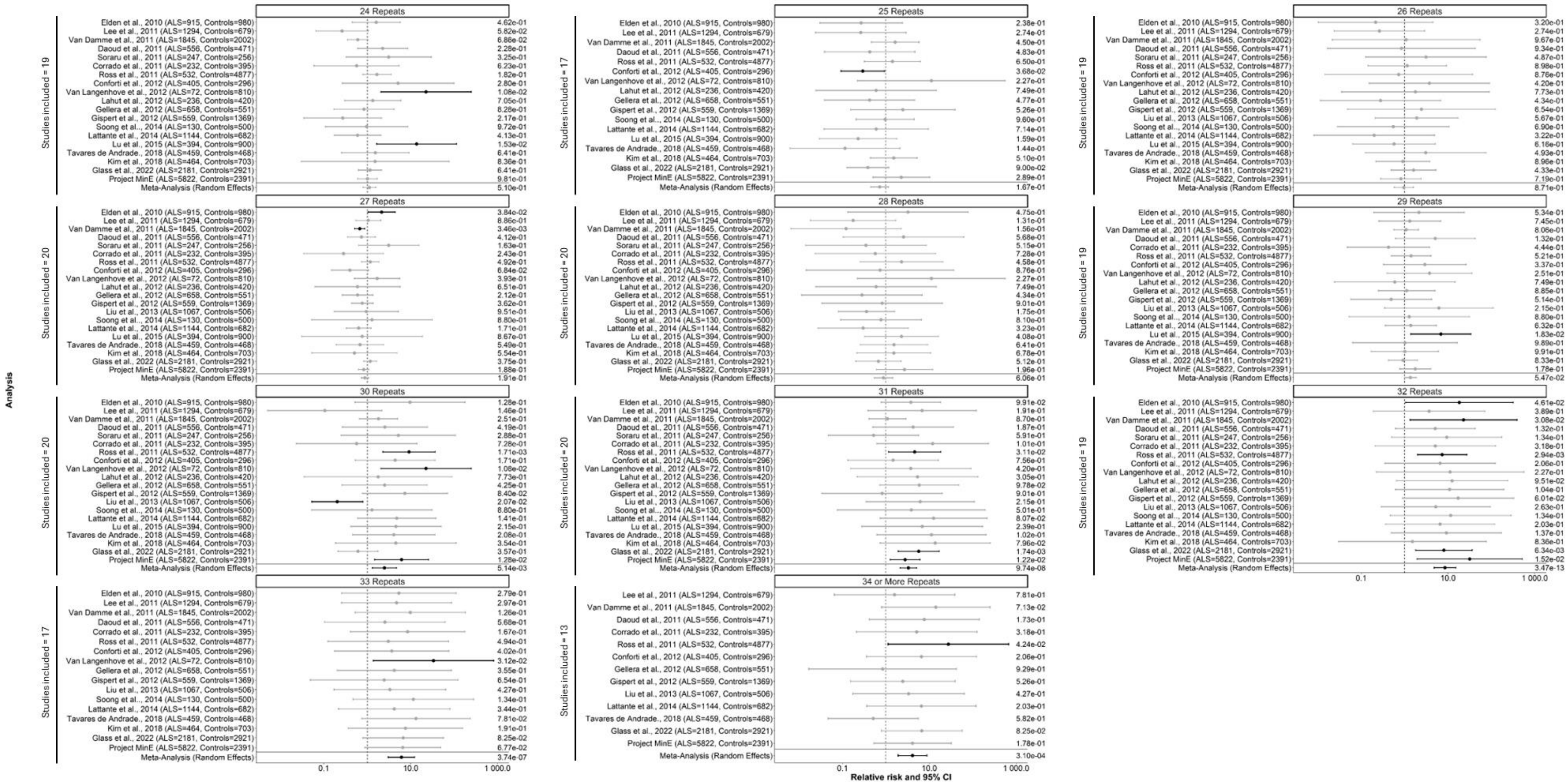

**Supplemental Figure 3. Meta-analysis of 19 studies presenting *ATXN2* repeat lengths of individuals with ALS and controls with the Project MinE Sequencing Consortium Dataset.** Relative risk and 95% confidence intervals were calculated for each of the 20 total cohorts, applying a continuity correction (0.5 to each cell) when counts were zero. The Wald Z-test was applied to each study to test the significance of associations. Pooled estimates were generated using a random effects model following assessment using the the I^2 statistic and Cochran's Q test. Studies were excluded from the analysis of a specific repeat length if there were no individuals with ALS or controls with repeats of that specific length.

**Supplemental Table 1. Statistical analyses presented within the currently available peer-reviewed literature presenting associations between *ATXN2* repeat lengths of various sizes and ALS risk.**

| Author | Year | Repeat Length LL | ALS Total (n) | Control Total (n) | ALS Carriers (n) | Control Carriers (n) | Odds Ratio | Lower CI | Upper CI | p-value | Statistical Test Applied | Population | Comments |
|---|---|---|---|---|---|---|---|---|---|---|---|---|---|
| Elden et al[2] | 2010 | 27 | 915 | 980 | 43 | 14 | 2.80 | 1.54 | 5.12 | 3.60E-05 | Unknown | White | First manuscript to propose association between *ATXN2* and ALS |
| Chen et al[30] | 2011 | 27 | 345 | 350 | 12 | 4 | 3.12* | 1.00 | 9.76 | 4.00E-02 | Chi-square | Chinese | Only assessed repeats of ≥27 |
| Corrado et al[8] | 2011 | 31 | 232 | 395 | 7 | 0 | 25.92* | 1.48 | 454.87 | 8.90E-04 | Fisher exact | Italian | Only assessed repeats of ≥31 |
| Daoud et al[29] | 2011 | 28 | 556 | 471 | 35 | 19 | 1.60* | 0.90 | 2.83 | 9.00E-02 | Fisher exact | French Canadian | Assessed multiple lower limits |
| Daoud et al[29] | 2011 | 29 | 556 | 471 | 25 | 4 | 5.50 | 1.90 | 15.90 | 2.40E-04 | Fisher exact | French Canadian | Assessed multiple lower limits |
| Lee et al[9] | 2011 | 31 | 1284 | 679 | 13 | 0 | 14.43* | 0.86 | 243.11 | 6.20E-03 | Fisher exact | European | Only assessed repeats of ≥31 |
| Ross et al[28] | 2011 | 27 | 532 | 4877 | 33 | 197 | 1.58 | 1.08 | 2.31 | 2.00E-02 | Logistic regression | White | Only assessed repeats of ≥27 and ≥31 |
| Ross et al[28] | 2011 | 31 | 532 | 4877 | 9 | 9 | 5.57 | 1.95 | 15.88 | 1.00E-03 | Logistic regression | White | Only assessed repeats of ≥27 and ≥31 |
| Van Damme et al[3] | 2011 | 29 | 1948 | 2002 | 28 | 16 | 1.92 | 1.04 | 3.64 | 3.60E-02 | Fisher exact | European | Assessed multiple lower limits through ROC |
| Conforti et al[32] | 2012 | 28 | 405 | 296 | 22 | 3 | 5.83 | 1.71 | 9.78 | 1.00E-03 | Fisher exact | Italian | Only assessed repeats of ≥28 |
| Gellera et al[10] | 2012 | 31 | 231 | 551 | 15 | 1 | 38.19* | 5.01 | 290.93 | 1.40E-03 | Fisher exact | Italian | Assessed multiple lower limits, but only with statistical approach for ≥31 |
| Gispert et al[31] | 2012 | 27 | 559 | 749 | 23 | 30 | 1.03* | 0.59 | 1.79 | 9.20E-01 | Chi-square | Central European | Assessed multiple lower limits |
| Gispert et al[31] | 2012 | 30 | 559 | 749 | 7 | 1 | 9.43 | 8.27 | 10.59 | 1.00E-02 | Chi-square | Central European | Assessed multiple lower limits |
| Lahut et al[11] | 2012 | 31 | 236 | 420 | 4 | 0 | 16.28* | 0.87 | 303.68 | 1.70E-02 | Fisher exact | Turkish | Only assessed repeats of ≥31 |
| Laffita-Mesa et al[33] | 2013 | 30 | NA | NA | NA | NA | 2.16 | 1.76 | 2.65 | 5.00E-04 | Meta-analysis | Meta Analysis | Meta-analysis assessing multiple lower limits |
| Laffita-Mesa et al[33] | 2013 | 32 | NA | NA | NA | NA | 2.62 | 2.23 | 3.09 | 5.00E-04 | Meta-analysis | Meta Analysis | Meta-analysis assessing multiple lower limits |
| Liu et al[12] | 2013 | 31 | 1067 | 506 | 17 | 0 | 16.88* | 1.01 | 281.19 | 4.00E-03 | Chi-square | Chinese | Only assessed repeats of ≥31 |
| Lattante et al[4] | 2014 | 29 | 1144 | 682 | 22 | 0 | 27.36* | 1.66 | 451.79 | 5.60E-05 | Fisher exact | French | Only assessed repeats of ≥29 |
| Wang et al[34] | 2014 | 30 | 9042 | 13116 | 154 | 46 | 3.93 | 2.49 | 6.20 | 1.00E-04 | Chi-square | Meta Analysis | Meta-analysis assessing multiple lower limits |
| Borghero et al[13] | 2015 | 31 | 375 | 247 | 4 | 0 | 6.00* | 0.32 | 111.87 | 1.00E-04 | Unknown | Italian | Only assessed repeats of ≥31 |

| | | | | | | | | | | | | | |
|---|---|---|---|---|---|---|---|---|---|---|---|---|---|
| Chio et al[36] | 2015 | 28 | 672 | 509 | 66 | 13 | 4.16* | 2.27 | 7.62 | 1.00E-04 | Fisher exact | Italian | Only assessed repeats of ≥28 |
| Lu et al[35] | 2015 | 27 | 379 | 900 | 11 | 6 | 4.45* | 1.64 | 12.13 | 2.00E-03 | Chi-square | Chinese | Only assessed repeats of ≥27 |
| Lysogorskaia et al[37] | 2015 | 28 | 208 | 149 | 10 | 6 | 3.36 | 1.10 | 10.40 | 2.60E-02 | Chi-square | Russian | Only assessed repeats of ≥28 |
| Narain et al[38] | 2017 | 27 | 131 | 127 | 6 | 2 | 3.00 | 0.59 | 15.15 | 3.00E-01 | Chi-square | Indian | Only assessed repeats of ≥27 |
| Sproviero et al[7] | 2017 | 29 | 10888 | 15463 | 245 | 113 | 3.10* | 2.48 | 3.88 | 6.20E-18 | Chi-square | Meta Analysis | Meta-analysis assessing multiple lower limits |
| Kim et al[39] | 2018 | 26 | 464 | 703 | 15 | 13 | 1.77 | 0.84 | 3.76 | 1.70E-01 | Fisher exact | Korean | Assessed multiple lower limits |
| Kim et al[39] | 2018 | 27 | 464 | 703 | 12 | 8 | 2.31 | 0.94 | 5.69 | 6.90E-02 | Fisher exact | Korean | Assessed multiple lower limits |
| Kim et al[39] | 2018 | 28 | 464 | 703 | 11 | 5 | 3.39 | 1.17 | 9.82 | 2.10E-02 | Fisher exact | Korean | Assessed multiple lower limits |
| Kim et al[39] | 2018 | 29 | 464 | 703 | 9 | 3 | 4.62 | 1.24 | 17.14 | 1.60E-02 | Fisher exact | Korean | Assessed multiple lower limits |
| Kim et al[39] | 2018 | 30 | 464 | 703 | 7 | 0 | 23.07* | 1.31 | 404.85 | 1.50E-03 | Fisher exact | Korean | Assessed multiple lower limits |
| Tavares de Andrade et al[40] | 2018 | 27 | 459 | 468 | 29 | 12 | 2.56 | 1.26 | 5.08 | 5.00E-03 | Logistic regression | Brazilian | Only assessed repeats of ≥27 |
| Glass et al[14] | 2022 | 31 | 2181 | 2921 | 39 | 8 | 6.93 | 3.19 | 15.02 | 9.50E-07 | Logistic regression | American | Assessed multiple repeat lengths individually, then only assessed repeats of ≥31 |
| Grassano et al[42] | 2022 | 30 | 1043 | 755 | 41 | Unknown | 2.84 | 1.45 | 5.57 | 2.30E-03 | Fisher exact | Italian | Only assessed repeats of ≥30 |
| Hou et al[5] | 2022 | 29 | 736 | 365 | 14 | 0 | 14.67* | 0.87 | 246.63 | 3.30E-02 | Fisher exact | Chinese | Only assessed repeats of 29-34 |
| Nagy et al[41] | 2022 | 24 | 153 | 195 | 28 | 18 | 2.01 | 1.07 | 3.79 | 3.50E-02 | Fisher exact or Chi-square | Central European | Only assessed repeats of ≥24 |
| Borrego-Hernandez et al[43] | 2024 | 27 | 620 | 362 | 56 | 13 | 2.67 | 1.47 | 4.88 | 6.00E-04 | Fisher exact | Spanish | Assessed multiple lower limits |
| Borrego-Hernandez et al[43] | 2024 | 28 | 620 | 362 | 49 | 10 | 3.02 | 1.55 | 6.31 | 5.00E-04 | Fisher exact | Spanish | Assessed multiple lower limits |
| Borrego-Hernandez et al[43] | 2024 | 29 | 620 | 362 | 25 | 2 | 7.56 | 2.10 | 32.55 | 5.00E-04 | Fisher exact | Spanish | Assessed multiple lower limits |
| Novy et al[6] | 2024 | 29 | 414 | 712 | 7 | 3 | 4.81 | 1.09 | 21.18 | 3.80E-02 | Logistic regression | Norwegian | Only assessed repeats of ≥29 |
| Demaegd et al[15] | 2025 | 31 | 2262 | 2425 | 57 | 9 | 6.90 | 3.43 | 14.05 | 1.00E-10 | Fisher exact | Unknown | Only assessed repeats of ≥31 in two populations |
| Demaegd et al[15] | 2025 | 31 | 5242 | 2425 | 108 | 9 | 5.60 | 2.86 | 11.17 | 6.00E-10 | Fisher exact | European | Only assessed repeats of ≥31 in two populations |

Of the 34 manuscripts found to present *ATXN2* repeat lengths in case-control cohorts, 30 presented results of statistical association analyses for repeats of at least one length threshold. P-values were pulled directly from the identified manuscripts. Odds ratios and confidence intervals were pulled directly from the identified manuscripts, where available, otherwise were calculated using the presented carrier counts and sample sizes.

*Odds ratio was not provided within the manuscript but was manually calculated using carrier counts.

Abbreviations: CI, confidence interval; LL, lower limit.

**Supplemental Table 2. Meta-analysis of 19 studies presenting *ATXN2* repeat lengths of individuals with ALS and controls with the Project MinE Sequencing Consortium Dataset.**

Relative risk and 95% confidence intervals were calculated for each of the 20 total cohorts, applying a continuity correction (0.5 to each cell) when counts were zero. The Wald Z-test was applied to each study to test the significance of associations. Pooled estimates were generated using a random effects model following assessment using the the I^2 statistic and Cochran's Q test. Studies were excluded from the analysis of a specific repeat length if there were no individuals with ALS or controls with repeats of that specific length.

[See "2026 01 08_Supplemental Table 3_Table of Meta-Analysis Results_ATXN2_Dilliott et al.xlsx"]
