## Supplementary material for "Assessment of *ATXN2* Repeat Expansion Length and Risk of ALS: A Meta-Analysis": Project MinE ALS Sequencing Consortium

***Project MinE ALS sequencing consortium***
*Philip Van Damme, Philippe Corcia, Patrick Vourc'h, Orla Hardiman, Russell McLaughin, Marc Gotkine, Jan H. Veldink, Leonard H. van den Berg, Mamede de Carvalho, Jesus S. Mora Pardina, Monica Povedano, Peter M. Andersen, Markus Weber, Nazli A. Başak, Ammar Al-Chalabi, Chris Shaw, Pamela J. Shaw, Karen E. Morrison, John E. Landers, Jonathan D. Glass*

***Emails:***
*Philip Van Damme,*

*Philippe Corcia,*

*Patrick Vourc'h,*

*Orla Hardiman,*

*Russell McLaughin,*

*Marc Gotkine,*

*Jan H. Veldink,*

*Leonard H. van den Berg,*

*Mamede de Carvalho,*

*Jesus S. Mora Pardina,*

*Monica Povedano,*

*Peter M. Andersen,*

*Markus Weber,*

*Nazli A. Başak,*

*Ammar Al-Chalabi,*

*Chris Shaw,*

*Pamela J. Shaw,*

*Karen E. Morrison,*

*John E. Landers,*

*Jonathan D. Glass,*
